# Artificial intelligence-enabled reconstruction of the right ventricular pressure curve using the peak pressure value: a proof-of-concept study

**DOI:** 10.1101/2024.01.22.24301598

**Authors:** Ádám Szijártó, Alina Nicoara, Mihai Podgoreanu, Márton Tokodi, Alexandra Fábián, Béla Merkely, András Sárkány, Zoltán Tősér, Bálint Lakatos, Attila Kovács

## Abstract

Conventional echocardiographic parameters of right ventricular (RV) function are heavily afterload-dependent. Therefore, the incorporation of the RV pressure curve may enable the formulation of new parameters that reflect intrinsic RV function more accurately. Accordingly, we sought to develop an artificial intelligence-based method that can reconstruct the RV pressure curve based on the peak RV pressure.

We invasively acquired RV pressure in 29 heart failure patients before and after the implantation of a left ventricular (LV) assist device. Using these tracings, we trained various machine learning models to reconstruct the RV pressure curve of the entire cardiac cycle solely based on the peak value of the invasively acquired curve. The best-performing model was compared with the performance of two other methods that estimated RV pressure curves based on a reference LV and RV pressure curve, respectively.

Among the evaluated algorithms, the multilayer perceptron (MLP) achieved the best performance with an R^2^ of 0.887 [0.834 – 0.941]. The RV and LV reference curve-based methods achieved R^2^ values of 0.879 [0.815 – 0.943] and 0.636 [0.500 – 0.771], respectively. The MLP and the RV reference curve-based estimation showed good agreement with the invasive RV pressure curves (mean bias: -0.38 mmHg and -0.73 mmHg, respectively), whereas the LV reference curve-based estimation exhibited a high mean bias (+3.93 mmHg).

The proposed method enables the reconstruction of the RV pressure curve, using only the peak value as input. Thus, it may serve as a fundamental component for developing new echocardiographic tools targeting the afterload-adjusted assessment of RV function.

**Graphical abstract:** *Key question*. Can artificial intelligence be useful in the reconstruction of the right ventricular pressure curve of the entire cardiac cycle using only the peak value as input? *Key findings*. Multilayer Perceptron predicted instantaneous pressure values with a balanced, low bias throughout the cardiac cycle, even slightly outperforming the reference curve creation methods. *Take-home message*. Accurate prediction of the right ventricular pressure curve may enable formulation of new echocardiographic parameters targeting the afterload-adjusted assessment of RV function.

LV: left ventricular, PVC: pulmonary valve closure, PVO: pulmonary valve opening, RV: right ventricular, TVC: tricuspid valve closure, TVO: tricuspid valve opening

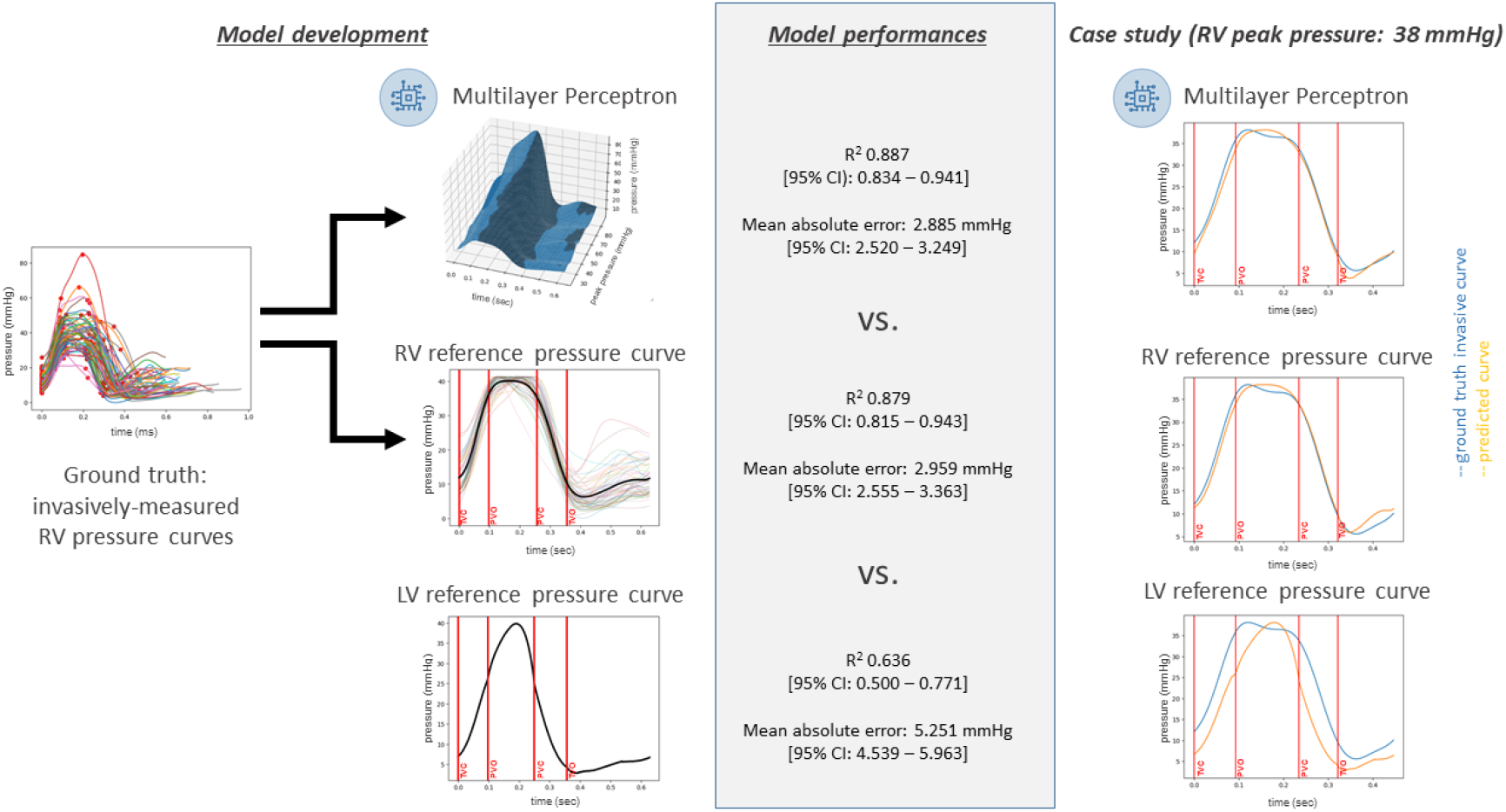

## Introduction

Echocardiography is the prime imaging modality to assess ventricular systolic function. However, the majority of the conventional parameters (*i*.*e*., ejection fraction - EF, global longitudinal strain - GLS) are heavily dependent on the actual afterload; thus, they reflect ventriculo-arterial coupling rather than intrinsic myocardial contractility.^1^ To create less load-dependent metrics that more accurately reflect myocardial contractility and energetics, the concept of myocardial work calculation has been recently introduced, and the added clinical value of the newly derived myocardial work parameters has been well established concerning the left ventricle (LV).^2–4^ A key step in myocardial work analysis is the non-invasive estimation of the LV pressure curve by rescaling a reference LV pressure curve (derived from LV pressure curves of several patients) based on a single, cuff-based measurement of systolic blood pressure. However, no dedicated solution exists for the right ventricle (RV) despite its function being even more dependent on afterload.^5^ Moreover, due to the different hemodynamic characteristics of the pulmonary vascular bed and the RV contraction pattern, the morphology of the RV pressure curve varies significantly between normal, mildly, or severely elevated pulmonary pressures.^6^ Thus, instead of deriving a single reference curve, a more sophisticated approach would be required that also considers the changes in the morphology of the RV pressure curve along the entire spectrum of peak RV pressures.

Accordingly, the aims of our proof-of-concept study were (1) to develop an artificial intelligence-based method that enables the estimation of the RV pressure curve based on the peak RV pressure value only and (2) to compare the reconstructed pressure curves with those created using other pre-existing approaches.

## Methods

End-stage heart failure patients over 18 years of age were enrolled in a prospective study and underwent durable LV assist device implantation at the Duke University Medical Center (patients provided written informed consent; IRB approval No. Pro00107652). Exclusion criteria were exchange of a previously implanted assist device, presence of a percutaneous RV assist device or extracorporeal membrane oxygenator, history of heart transplantation, prior or concomitant tricuspid valve repair/replacement, and contraindication for performing transesophageal echocardiography. In our current study, we analyzed 29 patients who also underwent pressure-conductance catheterization. After the induction of general anesthesia and the institution of mechanical ventilation, a 7Fr high-fidelity pressure-conductance catheter (CD Leycom, Zoetermeer, The Netherlands) that was connected to a signal processor with a dedicated software solution (Inca, CD Leycom) was advanced into the RV. The catheter was positioned in the RV apex under echocardiographic guidance and adjusted until signals derived from the individual electrode pairs were in phase. RV pressure tracings were obtained before skin incision (beginning-of-procedure examination) and after skin closure (end-of-procedure examination). A continuous pressure tracing containing 10 stable, separated cardiac cycles per examination was exported in a digital format with a 250Hz sampling rate (29 patients x 2 examinations x 10 cardiac cycles = a total of 580 cardiac cycles). Comprehensive 2D and 3D transesophageal echocardiography was performed during the operation to quantify RV volumes and RVEF (EPIQ CVx system, X8-2t transducer, Philips Healthcare, Best, The Netherlands; and 4D RV-Function 2, TomTec, Unterschleissheim, Germany).

The exported RV pressure tracings were processed using a custom software solution (implemented in Python v3.9) to identify dedicated cardiac cycle events (*i*.*e*., tricuspid valve closure – TVC, pulmonary valve opening – PVO, pulmonary valve closure – PVC, tricuspid valve opening – TVO) using the second derivative squared method by expert consensus reading.^7^ These annotated pressure tracings were split into segments, each containing the pressure curve of exactly one cardiac cycle (from TVC to TVC), which were then processed using a previously described method.^2^ Briefly, the curves were stretched or compressed along the time axis between each event to make the corresponding cardiac cycle events coincide and were scaled vertically (*i*.*e*., along the pressure axis) to have the same peak value. Then, the resulting normalized pressure curves from the same examination were averaged to create an examination-level pressure curve. The reference RV pressure curve was generated by normalizing and averaging all examination-level curves using the same method described above. Individual pressure curves were reconstructed by scaling this reference curve vertically based on the peak RV pressure and horizontally based on the timings of the cardiac cycle events. In the current study, we used the invasively measured, examination-level peak pressures to reconstruct the curves.

We have also evaluated the performance of the LV reference pressure curve-based estimation applied in a commercially available software solution (EchoPAC v204, GE Healthcare, Horten, Norway) for LV myocardial work calculation.

To enable a more accurate estimation of the RV pressure curve that respects the individual characteristics of the curve shape and dynamics along the scale of elevated pressures, we need to consider the effect of the peak RV pressure value. To achieve that, instead of simply averaging the temporally and vertically normalized curves, we opted for a machine learning-based method. Accordingly, we defined the task for the machine learning models to predict pressure values throughout the entire cardiac cycle for a given peak RV pressure value. In other words, the task was to fit a continuous 3D surface to all available examination-level pressure curves plotted as a function of the peak pressure values (Figure 1A). To reconstruct a patient’s pressure curve with a given peak RV pressure value, we need to run the model on all normalized time points, resulting in a temporally normalized curve. Again, we used the invasively measured, examination-level peak pressures for the reconstruction. Finally, the curve is denormalized along the time axis using the original timings of the cardiac cycle events. We experimented with several machine learning models (linear regression, K-nearest neighbors, support vector machines, random forest, multilayer perceptron – MLP). Leave-one-examination-out cross-validation was performed to quantify the models’ predictive accuracy (values averaged over all time points of the cardiac cycle), and the coefficient of determination (R^2^) was used as the performance metric. We also calculated the mean squared error (MSE) and the mean absolute error (MAE). The performance of the best-performing machine learning model was also compared with the performance of the LV and RV reference pressure curve-based estimation methods.

**Figure 1.**
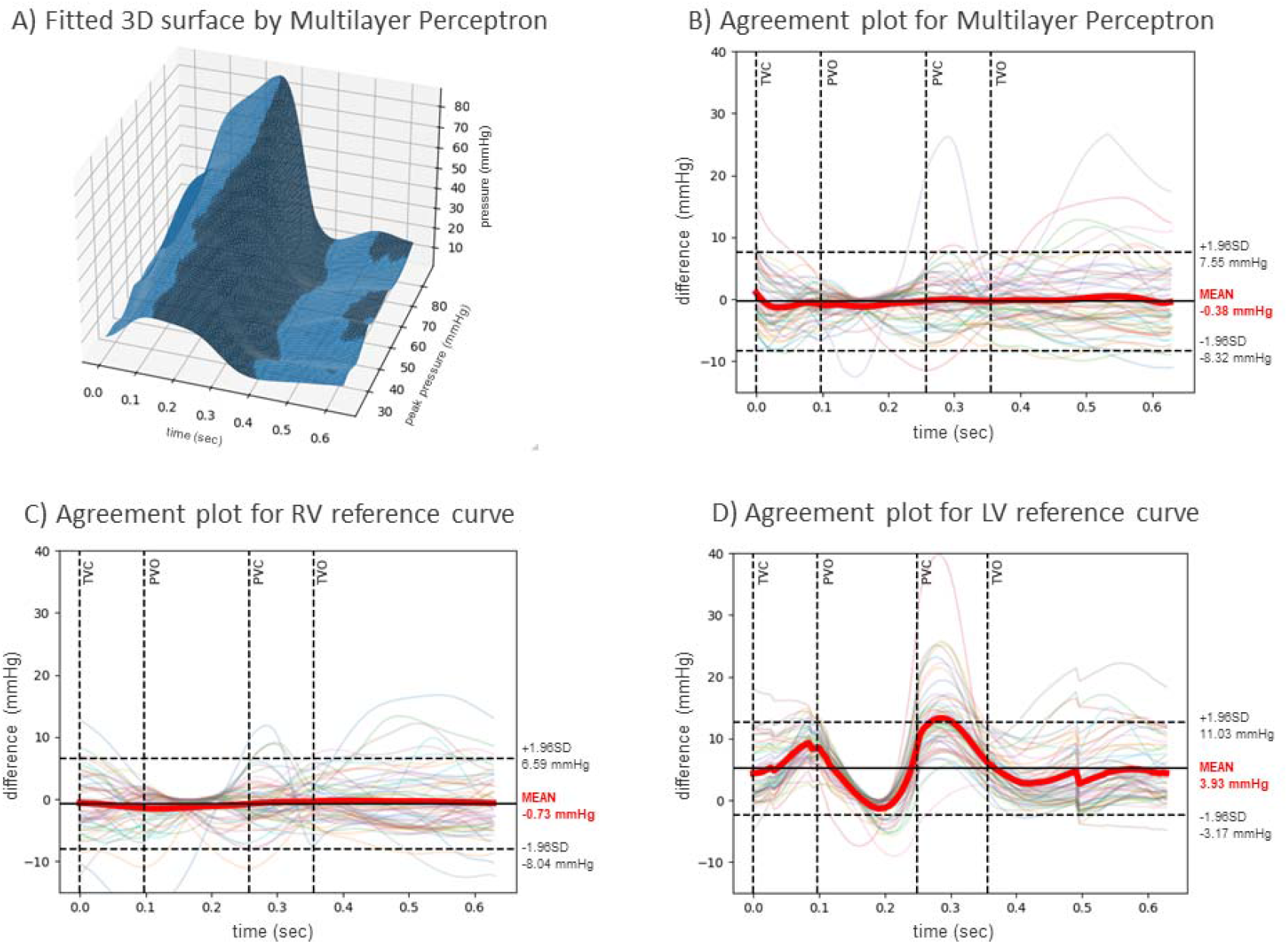
**(A)** The 3D continuous surface fitted by multilayer perceptron to the examination-level normalized RV pressure curves plotted as a function of the peak pressure values. **(B-D)** Agreement between the invasively measured ground truth and the pressure curves predicted by the three investigated methods for all examinations. The thick red line indicates the mean difference. LV: left ventricular, PVC: pulmonary valve closure, PVO: pulmonary valve opening, RV: right ventricular, TVC: tricuspid valve closure, TVO: tricuspid valve opening

Normal distribution of variables were tested using the Shapiro-Wilk test. Continuous variables are expressed as mean ± standard deviation or median (interquartile range) as appropriate, whereas categorical variables were reported as frequencies and percentages. Preoperative and postoperative values of echocardiographic measures were compared with paired t-test. Results from different pressure estimation methods were compared with the ground truth using Wilcoxon signed-rank tests and agreement plots. A two-sided p-value of < 0.05 was considered statistically significant.

## Results

Among the 29 patients undergoing LV assist device implantation, 22 (75.9%) were male, the mean age of the population was 61±14 years, and 10 patients (34.5%) were of nonischemic etiology. Before the implantation, 1 patient (3.4%) had trace, 18 (62.1%) had mild, 9 (31.0%) had moderate, and 1 (3.4%) had severe tricuspid regurgitation. RV end-diastolic (preop vs. postop, 168±58 vs. 140±49 mL, p<0.001) and end-systolic volumes (124±51 vs. 107±42 mL, p=0.01), along with RVEF (28±10 vs. 25±6%, p<0.05) decreased after the procedure. The mean RV peak systolic pressure of all examinations was 41.3±10.3 mmHg, ranging from 21.2 to 84.9 mmHg.

Among the evaluated machine learning algorithms, the MLP achieved the best performance with an R^2^ of 0.887 [95% confidence interval (CI): 0.834 – 0.941], an MSE of 14.398 [95% CI: 10.644 – 18.151] mmHg^2^, and an MAE of 2.885 [95% CI: 2.520 – 3.249] mmHg. The RV reference curve-based estimation method achieved an R^2^ of 0.879 [95% CI: 0.815 – 0.943], an MSE of 15.625 [95% CI: 10.919 – 20.329] mmHg^2^, and an MAE of 2.959 [95% CI: 2.555 – 3.363] mmHg, whereas the LV reference curve-based approach had an R^2^ of 0.636 [95% CI: 0.500 – 0.771], an MSE of 51.499 [95% CI: 37.875 – 65.124] mmHg^2^, and a MAE of 5.251 [95% CI: 4.539 – 5.963] mmHg. We investigated the differences between the invasively measured pressures and the predicted values at the valvular events (Table 1). For the MLP and the RV reference curve-based method, the estimated pressures did not differ significantly from the invasively measured ground truth values at TVC, PVC, and TVO (with the RV reference curve having a borderline non-significant overestimation at PVC). At PVO, both the MLP and the RV reference curve-based method significantly overestimated the RV pressure. The LV reference curve-based method significantly underestimated the pressure values at all dedicated time points. Agreement plots showed similar results (Figure 1B-D): whereas the MLP and the RV reference curve-based estimation had a low, balanced bias throughout the cardiac cycle (with the MLP having a slightly lower mean bias), the LV reference curve-based method showed the worst agreement with the ground truth.

**Table 1.**
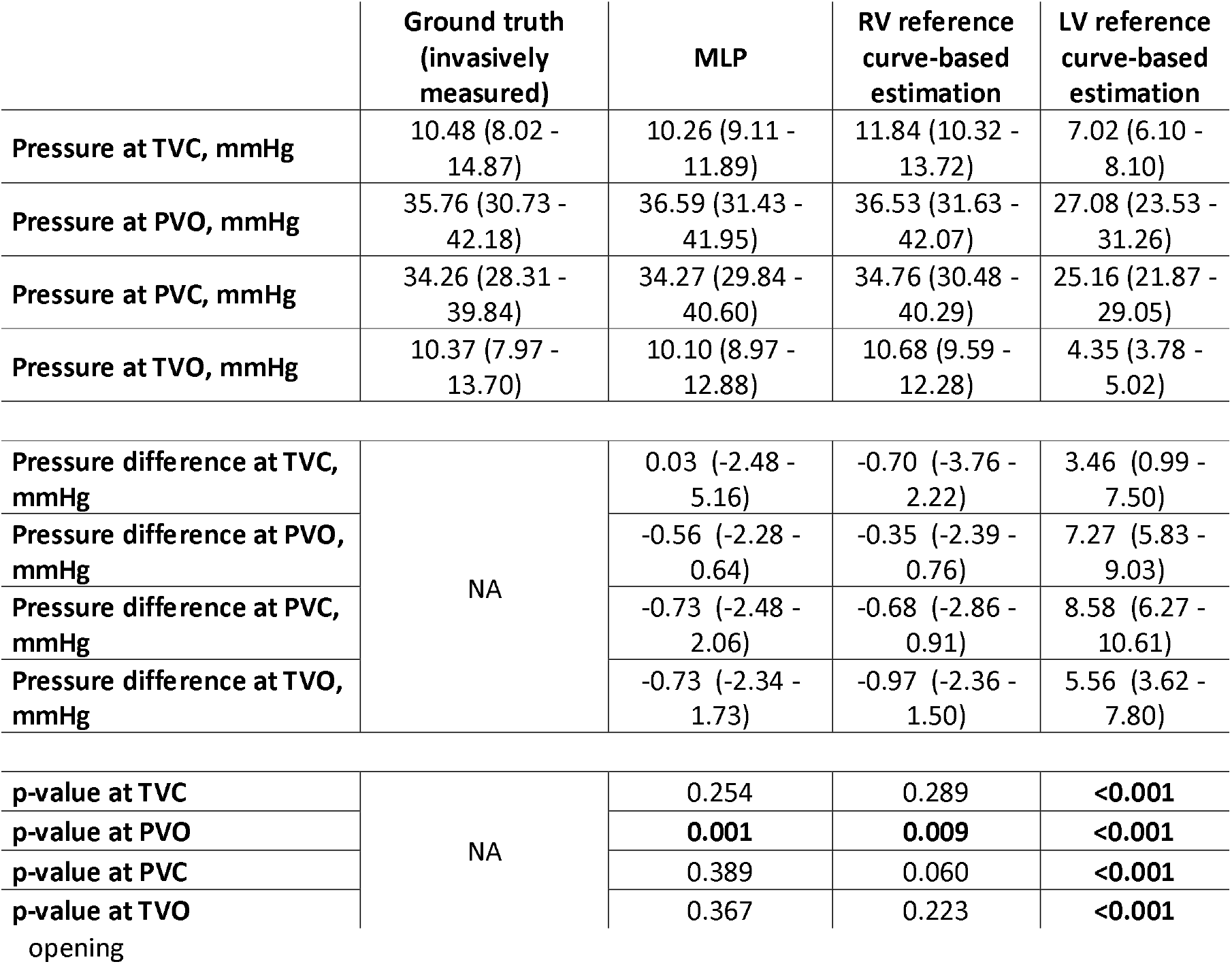
The invasively measured ground truth and the pressure values estimated by the three methods. LV: left ventricular, MLP: multilayer perceptron, PVC: pulmonary valve closure, PVO: pulmonary valve opening, RV: right ventricular, TVC: tricuspid valve closure, TVO: tricuspid valve

## Discussion

Our proof-of-concept study aimed to provide an artificial intelligence-based solution to reconstruct individual RV pressure curves with high fidelity using the peak pressure value as input. The proposed MLP showed promising results: it predicted RV pressure values with a balanced, low bias throughout the cardiac cycle, even slightly outperforming the RV reference curve-based method. As the morphology of the RV pressure curve can change markedly along the spectrum of elevated pressures (*i*.*e*., early systolic peaking transitions to a late peaking, notching pattern as pressure rises), a reconstruction that respects the actual peak RV pressure would be highly advantageous compared to a simple “averaging”. A physiologically more realistic curve for the given clinical scenario can have downstream consequences on RV function quantification by myocardial work or other methods relying on RV pressure curves.^8^ Doppler-based measurement of peak RV pressure is feasible using the jet of tricuspid or pulmonary valve regurgitation by echocardiography; thus, the proposed method can be easily translated to a widely available, non-invasive tool. It is important to emphasize that according to our results, the LV pressure reference curve-based estimation (and hence the commercially available dedicated LV solution) should not be used for RV myocardial work calculations, as it significantly underestimates RV pressures throughout the cardiac cycle.

Some limitations of our study have to be acknowledged. The performance of the proposed model is specific to the patient cohort from which it was derived; thus, RV pressure curves of patients with other disease etiologies should be incorporated to create a more comprehensive model. We hypothesize that by investigating more patients and various cardiopulmonary disease etiologies and stages, the added value of our method would be even more pronounced. Future studies should be conducted to validate our model externally and assess the impact of these differences in pressure curve reconstruction on the values of RV functional parameters (*e*.*g*., myocardial work indices).

## Data Availability

All data produced in the present study are available upon reasonable request to the authors

## Data availability

Data will be made available by the corresponding author for reasonable requests.

## References

1. Ruppert M, Lakatos BK, Braun S, et al. Longitudinal Strain Reflects Ventriculoarterial Coupling Rather Than Mere Contractility in Rat Models of Hemodynamic Overload-Induced Heart Failure. J Am Soc Echocardiogr 2020;33:1264–1275 e1264. 10.1016/j.echo.2020.05.017

2. Russell K, Eriksen M, Aaberge L, et al. A novel clinical method for quantification of regional left ventricular pressure-strain loop area: a non-invasive index of myocardial work. Eur Heart J 2012;33:724–733. 10.1093/eurheartj/ehs016

3. Lakatos BK, Ruppert M, Tokodi M, et al. Myocardial work index: a marker of left ventricular contractility in pressure-or volume overload-induced heart failure. ESC Heart Fail 2021;8:2220–2231. 10.1002/ehf2.13314

4. Tokodi M, Olah A, Fabian A, et al. Novel insights into the athlete’s heart: is myocardial work the new champion of systolic function? Eur Heart J Cardiovasc Imaging 2022;23:188–197. 10.1093/ehjci/jeab162

5. Kovacs A, Lakatos B, Tokodi M, Merkely B. Right ventricular mechanical pattern in health and disease: beyond longitudinal shortening. Heart Fail Rev 2019;24:511–520. 10.1007/s10741-019-09778-1

6. Richter MJ, Hsu S, Yogeswaran A, et al. Right ventricular pressure-volume loop shape and systolic pressure change in pulmonary hypertension. Am J Physiol Lung Cell Mol Physiol 2021;320:L715–L725. 10.1152/ajplung.00583.2020

7. Oakland H, Joseph P, Naeije R, et al. Arterial load and right ventricular-vascular coupling in pulmonary hypertension. J Appl Physiol (1985) 2021;131:424–433. 10.1152/japplphysiol.00204.2021

8. Richter MJ, Yogeswaran A, Husain-Syed F, et al. A novel non-invasive and echocardiography-derived method for quantification of right ventricular pressure-volume loops. Eur Heart J Cardiovasc Imaging 2022;23:498–507. 10.1093/ehjci/jeab038

